# Factors driving availability of COVID-19 convalescent plasma: Insights from a demand, production and supply model

**DOI:** 10.1101/2020.10.25.20219170

**Authors:** W. Alton Russell, Eduard Grebe, Brian Custer

## Abstract

**Background:** COVID-19 Convalescent Plasma (CCP) is a promising treatment for COVID-19. Blood collectors have rapidly scaled up collection and distribution programs.

**Methods:** We developed a detailed simulation model of CCP donor recruitment, collection, production and distribution processes. Simulations based on epidemics in 11 U.S. states, in which key parameters were varied over wide ranges, allowed identification of the drivers of ability to calibrate collections capacity and ability to meet demand for CCP.

**Results:** Changes in collection capacity utilization lagged increases and decreases in COVID-19 hospital discharges, and never exceeded 75% in most simulations. Demand could be met for most of the simulation period in most simulations, but in states with early sharp increases in hospitalizations a substantial portion of demand went unmet during these early peaks. Modeled second wave demand could generally be met with stockpiles established during first epidemic peaks. Apheresis machine capacity (number of machines) and probability that COVID-19 recovered individuals are willing to donate were the most important supply-side drivers of ability to meet demand. Recruitment capacity was important in states with early peaks.

**Conclusions:** Epidemic trajectory was the most important determinant of ability to meet demand for CCP, although our simulations revealed several contributing operational drivers of CCP program success.

## INTRODUCTION

The novel coronavirus SARS-CoV-2 has fueled a global pandemic, with more than 37 million confirmed COVID-19 cases and 1 million deaths as of October 12, 2020 [1]. Transfusion of convalescent plasma from recovered individuals with a mature antibody response has been successfully used for post-exposure prophylaxis and treatment during other disease outbreaks, including two other coronaviruses: severe acute respiratory syndrome (SARS-1) and Middle East respiratory syndrome (MERS) [2,3]. In response to the SARS-CoV-2 pandemic, many countries and blood collection agencies rapidly established COVID-19 Convalescent Plasma (CCP) collection, processing, and distribution programs during the first half of 2020, with several clinical trials underway. Preliminary evidence suggests that CCP is a safe treatment for COVID-19 [4], and ongoing randomized controlled trials are evaluating its efficacy [5]. Because CCP is a new and unique blood product, limited data are available regarding the operational challenges of CCP collection and distribution programs during an epidemic.

Using data from Vitalant, a nonprofit that collects approximately 14% of the US blood supply, we developed a simulation model of CCP donor recruitment, donation collection, testing, and distribution processes. In this paper, we use our simulation to evaluate how epidemic trajectory, donor recruitment and retention, collections capacity, and demand impact ability to meet competing priorities of current clinical demand and stockpiling CCP for future use in 11 different U.S. states. Our aim was not to replicate CCP collections in each state but rather to analyze and gain insights into the diverse set of drivers of a successful CCP program.

## METHODS

We ran our simulation for eleven epidemic trajectories while varying parameters related to CCP donor recruitment and return, collections capacity, and demand. Epidemic trajectories were based on calibrated state-level SEIR epidemic models developed and published by ‘COVID Act Now’ [6]. We included 10 states with the highest cumulative per-capita COVID-19 hospitalization rate as of August 31, 2020 (New Jersey, New York, Massachusetts, Illinois, Louisiana, Connecticut, Indiana, Mississippi, Virginia, and Maryland) and California, which had the highest overall number of COVID-19 hospitalizations during this period. We excluded Washington D.C. despite having the highest estimated per-capita hospitalization rate because it had fewer than 10,000 total hospitalizations.

For each state we estimated daily hospital admissions and discharges from three reported outcomes in the COVID Act Now state-level models (hospital beds required, deaths, and infections by day) using a method described in the appendix. We ran 10,000 simulations of a 200-day period beginning on the date of the first discharge of a COVID-19 patient and calculated two daily outcomes: (1) the percent of CCP collection capacity utilized and (2) the percent of CCP demand unmet. In each simulation, we sampled seven input parameters related to donor recruitment and return, collections capacity, and CCP demand using uniform distributions.

Our simulation model consists of two linked components: (1) a microsimulation of the donor recruitment, return, collections and CCP screening processes and outputs a number of usable units collected by day (2) a production model that accounts for production lag, demand, distribution and inventory.

### Donor recruitment and return

In the simulation, potential CCP donors (agents) entered the model at discharge from hospital and become eligible for CCP donation 14 days later. Across simulations, we varied the probability a recovered individual would be willing to donate CCP from 10% to 90%, and we varied capacity for new donor recruitment from 0.2 to 2 new donors each day per apheresis machine. Willing donors who were recruited each day entered the donor pool. Each day, donors in the pool would be selected randomly and scheduled for donations, subject to a collection capacity. Scheduled donations could be incomplete due to donor deferral for other reasons (2% probability) or failed or incomplete donation (1% probability). Completed donations could be removed from the CCP supply due to testing positive for disease markers (0.2% probability), not meeting the SARS-CoV-2 antibody release criterion (8% probability), or testing positive for HLA antibodies (9% probability for female donors). These probabilities were based on data from the first seven months of Vitalant’s CCP program. Donors were eligible for another plasmapheresis donation after 7 days and remained eligible until 180 days post-discharge. For each donor, we sampled the minimum number of days until a subsequent donation from an empirical distribution fit to Vitalant CCP donation data. We developed separate distributions for time to return for the donor’s second, third, and fourth-or-greater donation, respectively, based on differences in return donation propensity observed in the data (Figure S1). We assumed that donors who do not return by 130 days would never return: 57%, 31%, and 11% of donors never returned for a second, third, or fourth-or-greater donation, respectively.

### Capacity

In each state we varied the per capita number of apheresis machines from 4 to 55 per million residents. We assumed that each machine could support on average 3.5 CCP collection procedures per day and that 50% of machine capacity would be available for CCP collections. We based the range of per-capita apheresis machines on three states where over 90% of apheresis donations are collected by Vitalant: Colorado (8 machines per million residents), South Dakota (34 machines per million residents), and North Dakota (55 machines per million residents).

### Demand

We calculated daily CCP demand from the estimated daily hospital admissions and 3 parameters: the probability each hospitalized COVID-19 patient requires CCP (varied from 10% to 45% of patients), the number of CCP units required per patient (varied from 1.5 to 4 units), and the average delay from admission to receiving CCP transfusion (varied from 1 to 5 days). In the simulation, the daily number of CCP units produced and any existing inventory was used to meet demand. If the available CCP exceeded demand it was added to inventory and available to meet future demand.

### Sensitivity analysis

For sensitivity analysis, we performed a separate set of simulations in which we also varied donor return. To do so, we fit an exponential distribution to the probability a donor returns by *t* days from their last donation of the form

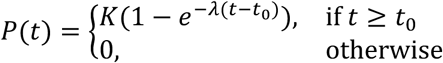

We set *t*_0_ (the minimal return time) to 7 days in line with current practice, and fit *λ* (the exponential rate parameter) and *K* (the asymptote limiting donor return) to Vitalant data using maximum likelihood estimation. As before, we used separate distributions for the second, third, and fourth-or-greater donations. To vary return time, we added a scaling factor *s* as follows:

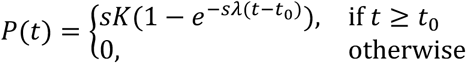

This distribution leads to greater return when *s* > 1 and less donor return when *s* < 1. In sensitivity analysis we varried *s* from 0.5 to 2.25 only in the distribution for second donations.

To assess the sensitivity of the total percent demand unmet to each parameter, we regressed each parameter on the outcome using locally estimated scatterplot smoothing (LOESS) regression for each state. We then predicted the outcome at the 1st, 25th, 50th, 75th, and 99th percentile of each input. We assessed the degree to which the predicted outcome changed depending on the quantile of parameter used to predict it, an indication of how sensitive the ability to meet demand was to the parameter (or how important the parameter was). We developed an easy-to-use web-based modeling tool, available at https://vitalantri.shinyapps.io/ccp_model. All code is open-source and publicly available [7].

## RESULTS

Across states, increases in collection capacity utilization lagged behind increases in COVID-19 patient discharge by 2 weeks, reflecting the delay in donor eligibility (Figure 1). In periods when discharges fell sharply, capacity utilization fell more gradually. In more than 50% of simulations across all states, percent capacity utilized never exceeded 75% of available machine time, indicating that fully utilizing available collection capacity may be an important challenge.

**Figure 1.**
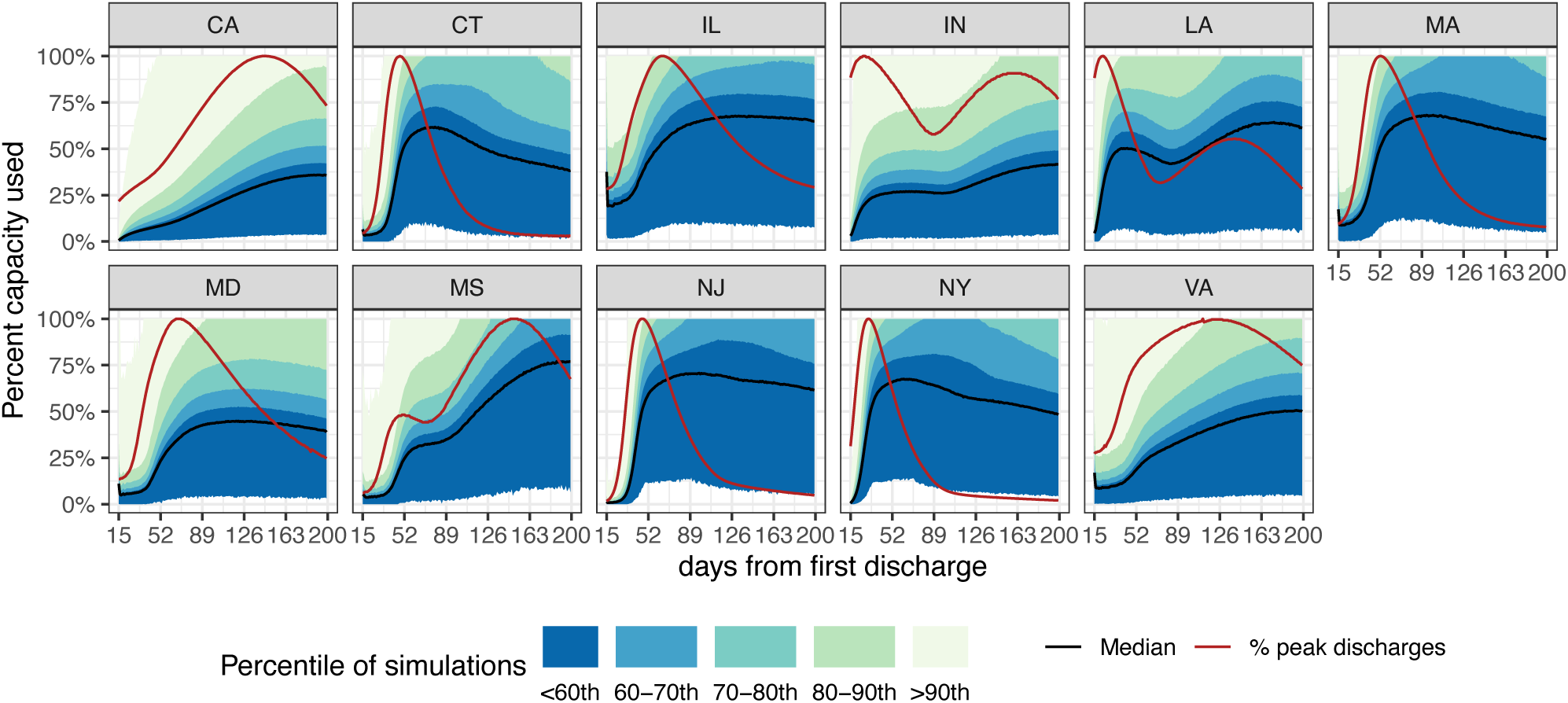
Percentiles of the percent capacity used by simulation day for each state across 10,000 simulations. Epidemic trajectories for each state shown by the percent of peak discharges by day plotted in red.

In most simulations, states met most of the demand for most of the period despite relatively low capacity utilization (Figure 2). Demand was more likely to go unmet during early increases in hospitalizations, particularly in states with very steep hospitalization increases (e.g., New York). Most of the demand was met in most simulation for states with more gradual increases (e.g., Virginia and California). In states with two epidemic peaks (e.g., Louisiana, Indiana) demand was mostly met during the second wave using inventory stockpiled during the downswing from the first peak.

**Figure 2.**
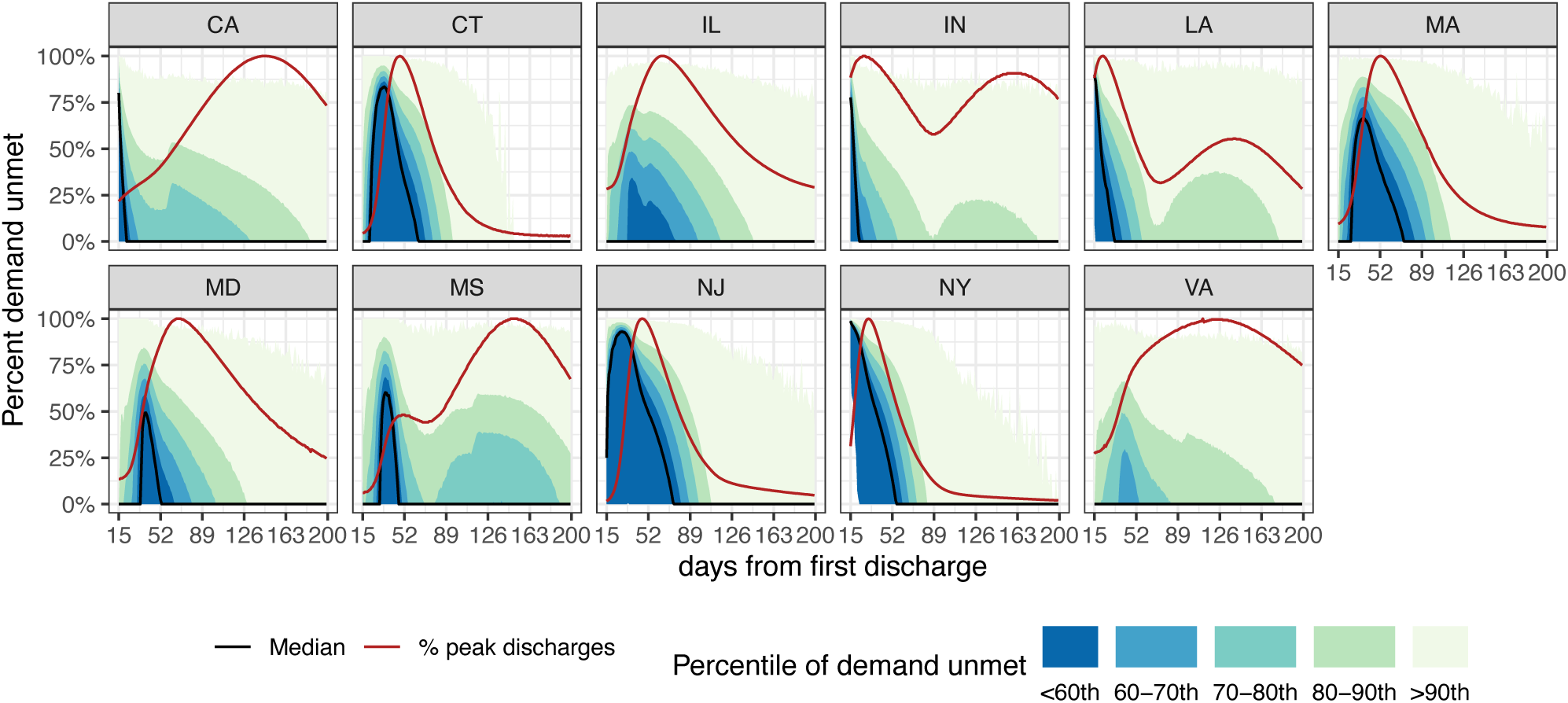
Percentiles of the percent demand unmet by simulation day for each state across 10,000 simulations. Epidemic trajectories for each state shown by the percent of peak discharges by day plotted in red.

The sensitivity of percent demand unmet to uncertain parameters is plotted in Figure 3 for four states and in Figure S2 for the other states. Apheresis machine capacity was a strong determinant of the percent demand unmet. Given that we assumed recruitment capacity was proportional to the number of machines, some of that benefit could be attributed to increased donor recruitment. Both the fraction of hospitalized patients requiring CCP and the number of units transfused per patient were more important parameters than the delay from admission to CCP administration. The scale multiplier on donor return did not substantially impact ability to meet demand. In most states the probability COVID-19 recovered individuals were willing to donate was the most important donor-related parameter. However, the daily donor recruitment capacity was more important in New York and New Jersey. Both these states experienced very early, steep epidemic trajectories that yielded many potential CCP donors, which may explain why capacity to recruit was a more important parameter than willingness to donate.

**Figure 3.**
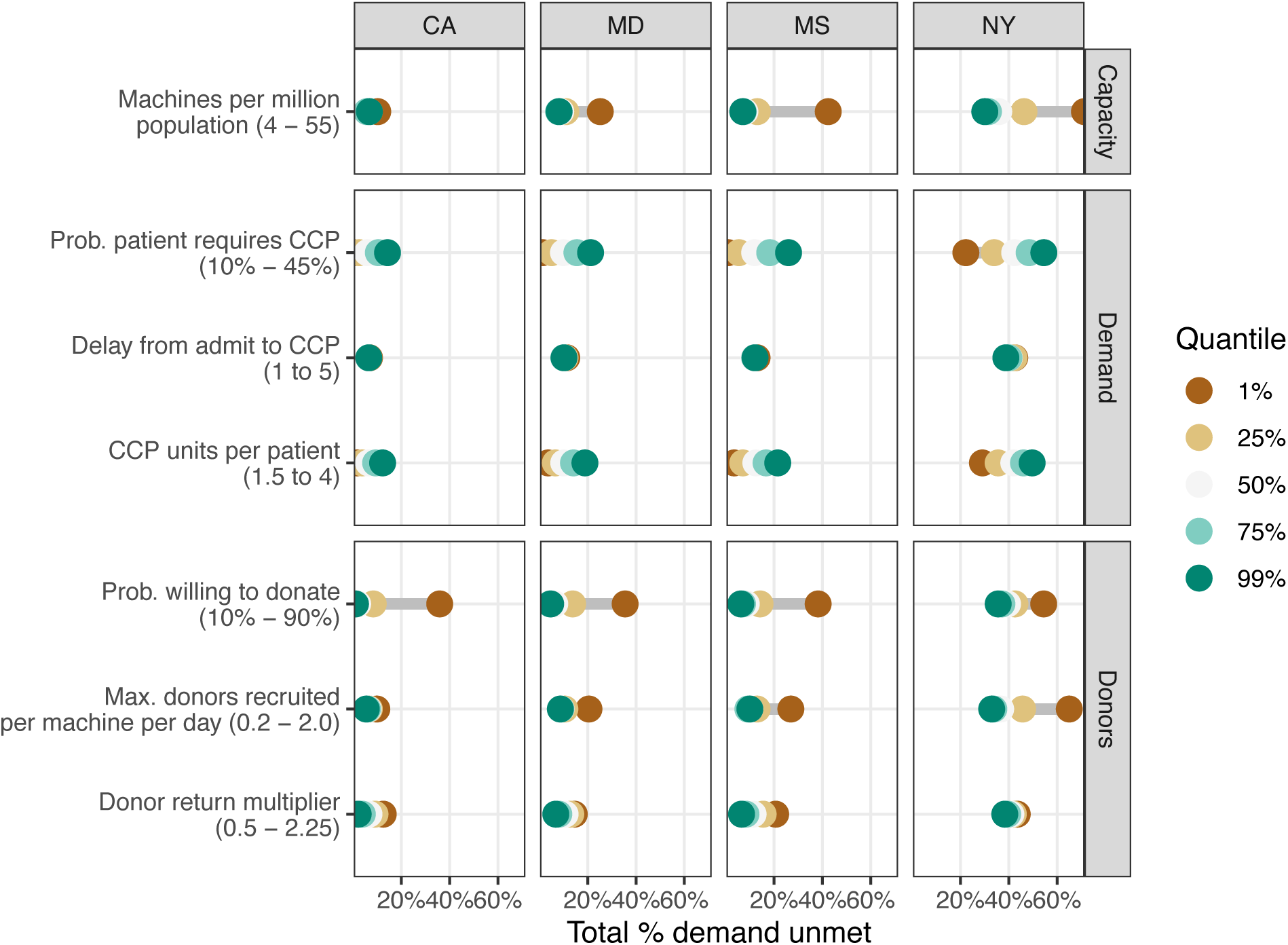
Sensitivity of percent demand unmet over simulation period to seven parameters. Four states with different epidemic trajectories shown here; remaining seven states are shown in appendix figure S2. The predicted percent demand unmet based on five quantiles of each parameter is plotted. Greater distance between plotted points indicates greater sensitivity to that parameter.

## DISCUSSION

While donor willingness to return was not a driver of outcomes in our simulations, the percentage of donors deferred or unwilling to return increased to very high levels over the simulation period in all states (Figure S3), indicating that it could be important over a longer time horizon and when insufficient CCP donors are recruited during initial epidemic peaks.

Unexpectedly, our analysis suggests that blood centers may struggle to fully utilize capacity and meet demand for CCP collections, particularly during rapid increases in the epidemic. In periods when demand is not quickly growing, blood centers can likely meet demand even when capacity utilization is fairly low. While ability to meet demand was sensitive to several parameters, epidemic trajectory was most important (Figure S4). Our simulations also demonstrated that having inventory on hand before an increase in demand greatly increased ability to meet demand.

The evidence base for CCP, and both operational practices and regulatory policies are evolving rapidly. Our simulations were informed by Vitalant’s CCP program, which may differ from that of other blood collectors. We further assumed an early start to and stable efforts in recruitment and collection, rather than a gradual ramp-up. Despite these limitations, our analysis reveals key drivers of the ability to utilize capacity and meet demand for CCP during an epidemic.

## Data Availability

We have shared all code and data in a public repository except for the individual donor return data. https://doi.org/10.5281/zenodo.4082755.

https://github.com/vitalantri/ccp_model

## APPENDIX

Eduard Grebe, W. Alton Russell, Brian Custer

## PROCEDURE FOR ESTIMATING DAILY HOSPITAL ADMISSIONS AND DISCHARGES FROM COVID ACT NOW MODEL OUTPUT

Our microsimulation model of the donor recruitment and collection process relies on a virtual cohort of potential CCP donors. At each time step, new agents are created equal to the estimated number of individuals discharged from hospital after recovery from severe COVID-19. Our production and supply model relies on the estimated number of new ICU admissions by day. We estimated the number of discharges from hospital and admissions to ICU from the output of state-level SEIR epidemic models published by ‘COVID Act Now’.

The published ‘COVID Act Now’ model output time series do not include new recoveries or hospital admissions and discharges. However, these can be estimated from the variables that are provided:

- cumulativeDeaths (*cD*_*t*_)
- cumulativeInfected (*cI*_*t*_)
- currentInfected (*I*_*t*_)
- currentSusceptible (*S*_*t*_)
- currentExposed (*E*_*t*_)
- hospitalBedsRequired (*H*_*t*_)

Given the following identities:

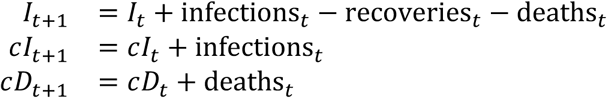

we can obtain the number of recoveries at each timestep by rearranging and substituting:

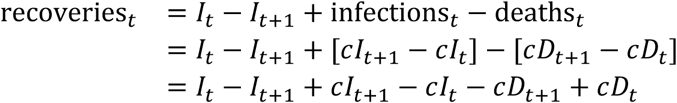

By assuming that the proportion of recoveries that represent discharges from hospital is equal to the proportion of infected individuals that are hospitalized, we can obtain the number of discharges from hospital by day:

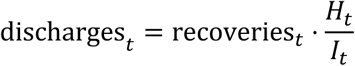

Once we have estimated the number of discharges by day, we are able to obtain the number of new hospital admissions by day:

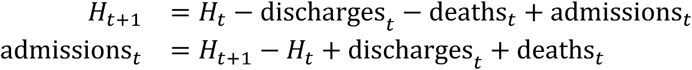

To estimate the number of ICU admissions by day, we assumed that:

- a certain proportion of patients hospitalized with COVID-19 never require critical care
- a certain proportion are admitted directly to the ICU (*p*_*d*_)
- a certain proportion are stepped up to critical care (*p*_*s*_) after a certain time (*τ*) in standard care

We assumed *p*_*d*_ = 0.04 and *p*_*s*_ = 0.12, which are similar to observed values in hospitalization data from Canada. The distribution of times from admission to care step-up has a long tail, but the vast majority of patients who are stepped up to critical care are admitted to the ICU in the first few days after hospitalization, with a median delay of 2 days (*τ* = *c*). For the purposes of estimating ICU admissions by day we used the median value.

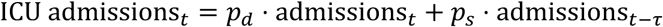

## DECLARATIONS

### Ethics/Consent

No human subjects or identifiable data were involved in this analysis.

### Data and materials

We have shared all data in a public repository except for the individual donor return data.

### Code availability

We have shared all code in a public repository (https://doi.org/10.5281/zenodo.4082755).

### Authors’ contributions

All authors developed the research question and methods and critically reviewed the manuscript. All authors identified sources and obtained data for the model. E.G. and W.A.R. conducted the analysis (E.G. led development of the donor microsimulation and W.A.R. led development of the policy analysis) and wrote the manuscript. B.C. provided critical review of the manuscript.

## Acknowledgements

The authors thank the many Vitalant team members who provided expertise and data that informed model development. The authors also thank the COVID Act Now team for developing the epidemic model used for this analysis (http://covidactnow.org/).

---

**Figure S1.**
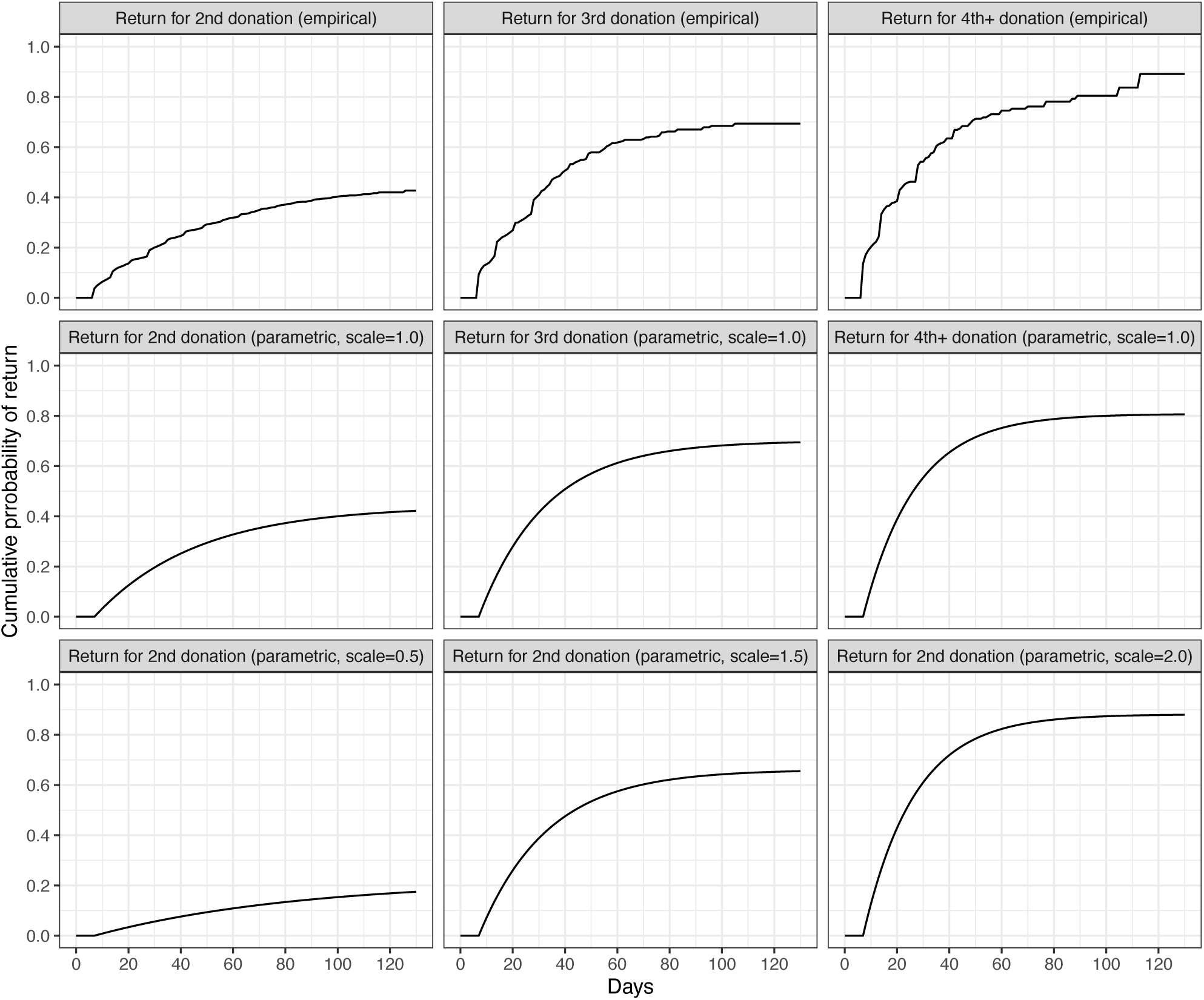
Empirical donor return distributions used in primary analysis (top row), parametric versions with scaling factor set to 1 (middle row), and scaled parametric distributions used in sensitivity analysis (bottom row)

**Figure S2.**
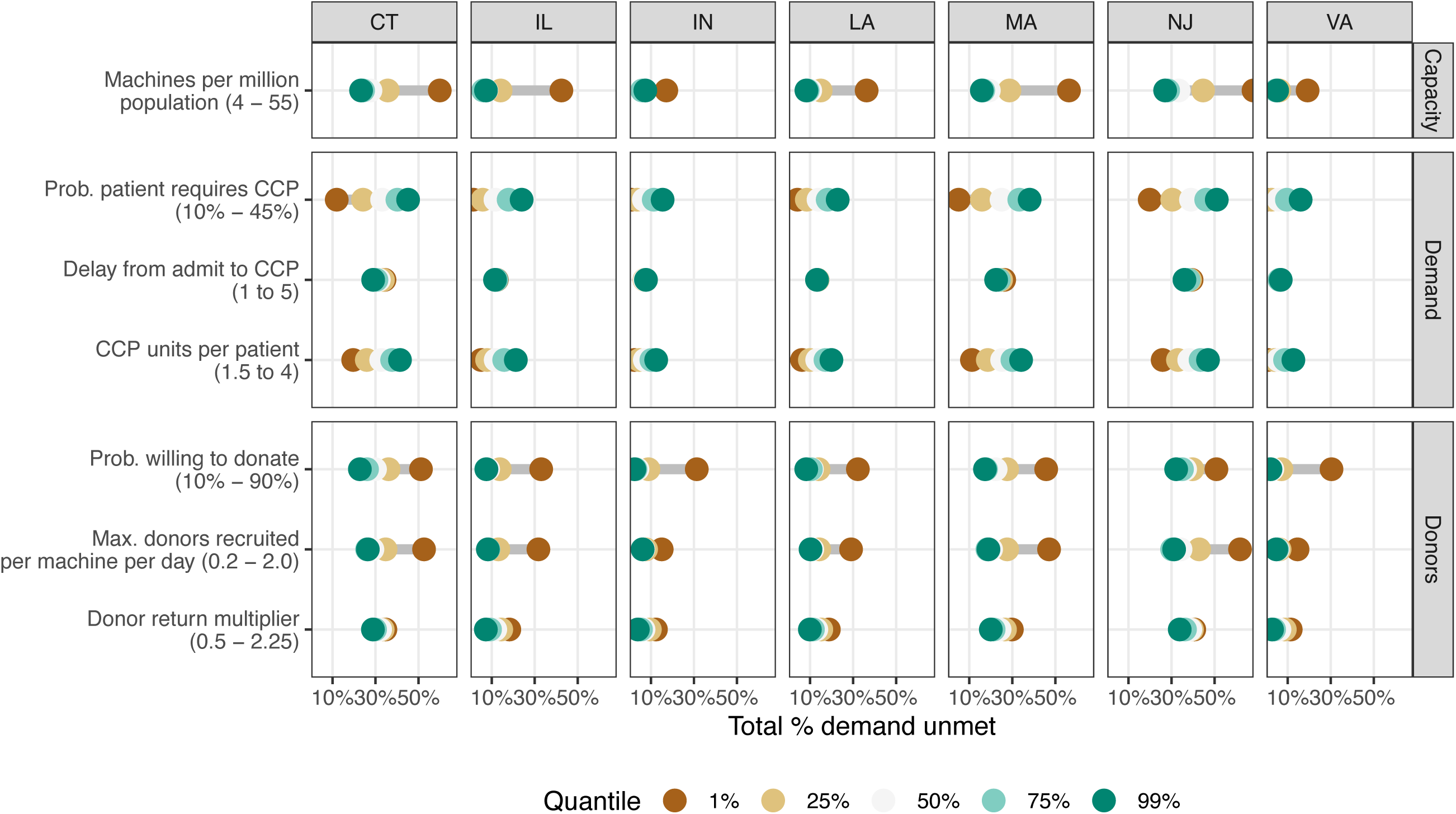
Sensitivity of percent demand unmet over simulation period to seven parameters. Seven states shown here; other four states are shown in Figure 3. The predicted percent demand unmet based on five quantiles of each parameter is plotted. Greater distance between plotted points indicates greater sensitivity to that parameter.

**Figure S3.**
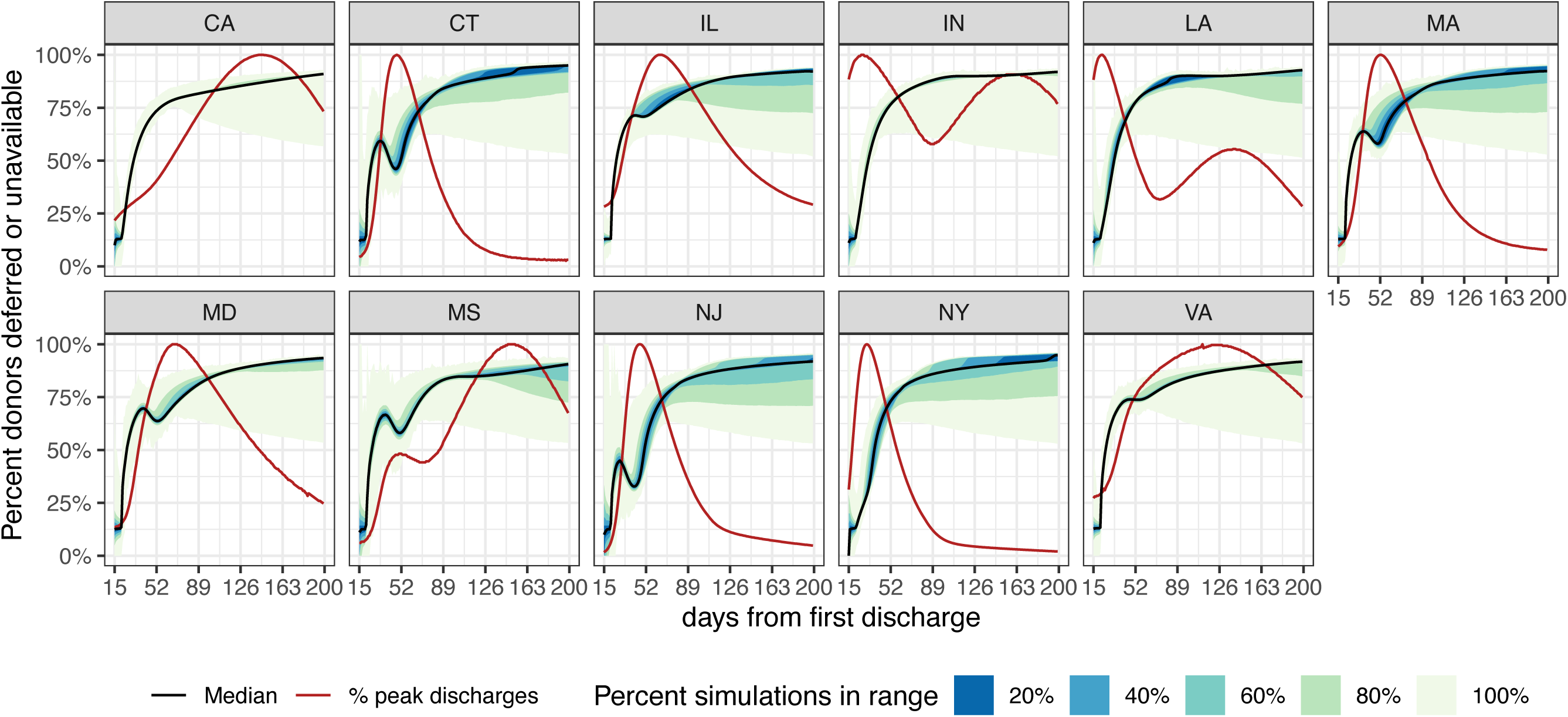
Donors deferred or unwilling to return over time by state. Over longer time horizons donor willingness to return may be a significant constraint on collections.

**Figure S4.**
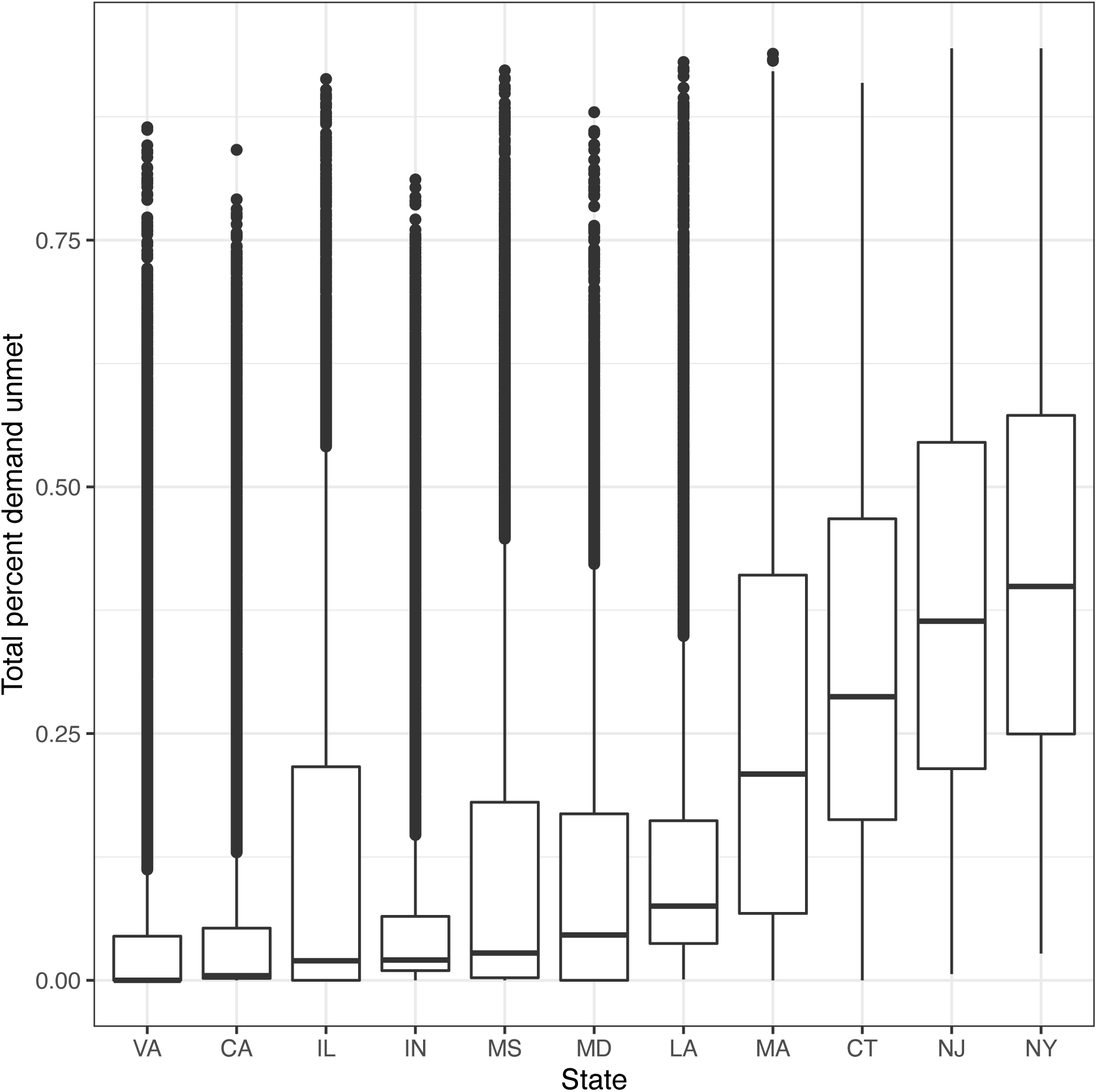
Boxplots showing the distribution of total percent demand unmet by state. The states that failed to meet the most demand experienced sharp, early increases in COVID-19 hospitalizations.

